# multiSero: open multiplex-ELISA platform for analyzing antibody responses to SARS-CoV-2 infection

**DOI:** 10.1101/2021.05.07.21249238

**Authors:** Janie R. Byrum, Eric Waltari, Owen Janson, Syuan-Ming Guo, Jenny Folkesson, Bryant B. Chhun, Joanna Vinden, Ivan E. Ivanov, Marcus L. Forst, Hongquan Li, Adam G. Larson, Wesley Wu, Cristina M. Tato, Krista M. McCutcheon, Michael J. Peluso, Timothy J. Henrich, Steven G. Deeks, Manu Prakash, Bryan Greenhouse, John E. Pak, Shalin B. Mehta

## Abstract

Serology has provided valuable diagnostic and epidemiological data on antibody responses to SARS-CoV-2 in diverse patient cohorts. Deployment of high content, multiplex serology platforms across the world, including in low and medium income countries, can accelerate longitudinal epidemiological surveys. Here we report multiSero, an open platform to enable multiplex serology with up to 48 antigens in a 96-well format. The platform consists of three components: ELISA-array of printed proteins, a commercial or home-built plate reader, and modular python software for automated analysis (pysero). We validate the platform by comparing antibody titers against the SARS-CoV-2 Spike, receptor binding domain (RBD), and nucleocapsid (N) in 114 sera from COVID-19 positive individuals and 87 pre-pandemic COVID-19 negative sera. We report data with both a commercial plate reader and an inexpensive, open plate reader (nautilus). Receiver operating characteristic (ROC) analysis of classification with single antigens shows that Spike and RBD classify positive and negative sera with the highest sensitivity at a given specificity. The platform distinguished positive sera from negative sera when the reactivity of the sera was equivalent to the binding of 1 ng mL^−1^ RBD-specific monoclonal antibody. We developed normalization and classification methods to pool antibody responses from multiple antigens and multiple experiments. Our results demonstrate a performant and accessible pipeline for multiplexed ELISA ready for multiple applications, including serosurveillance, identification of viral proteins that elicit antibody responses, differential diagnosis of circulating pathogens, and immune responses to vaccines.

## Introduction

The 2019-2020 coronavirus disease (COVID-19) pandemic caused by the novel coronavirus SARS-CoV-2 has driven the design of numerous serological tests for antibodies against the virus. These tests have been most useful in epidemiological surveys that track the geographic and demographic distribution of virus infections (1–5). These assays have also been informative in estimating the prevalence of infection when real-time RT-PCR testing is not readily available and in identifying vulnerable populations (6). Despite the epidemiological value, serological testing is currently limited in low- and medium-income countries as shown by SeroTracker, an aggregator of global serosurveillance data on SARS-CoV-2 (7). Many of the high-performing commercially available serological assays require proprietary instruments to read the assay, and the cost of consumables, instruments, and analysis software can prohibit epidemiology surveys in resource-limited settings.

Relative to a single antigen serological test, a multiplex (i.e., a multi-antigen) serological test can provide the following advantages: a) simultaneous interpretation of the magnitude of response to multiple viral antigens and vaccine components (8–11), b) differential diagnosis of infection (12, 13), c) robust and quantitative measurement of antibody response, and d) increase in sensitivity due to greater coverage of immunogenic epitopes (13–19). The increased specificity and sensitivity of printed 3 multiplex ELISA are particularly valuable when the assay is used for patient populations and cohorts with varying severity of COVID-19 or time since infection, as mild and asymptomatic infections are associated with lower antibody titers compared to severe cases (20, 21) and antibody response may differentially wane depending on the cognate antigen (22). A panel of multiple SARS-CoV-2 antigens can also provide longitudinal measurement of antibody responses to various antigens within individuals. Longitudinal evaluation of responses to multiple antigens will be helpful in understanding which antibody responses most strongly associated with immunity to SARS-CoV-2, and how durable these responses are after infection or vaccination. In particular, multiplex serological tests will be important after widespread vaccine distribution to differentiate vaccine-induced responses from those induced by naturally acquired infection. Beyond the current COVID-19 pandemic, multiplex serological testing can be adapted for other endemic infectious agents as a tool for serosurveillance of pathogens and vaccine coverage (23).

Multiplexed ELISA follows an experimental workflow similar to conventional ELISA. Both assays consist of coating a plate with antigen(s), blocking, and overlaying an antibody-containing solution (e.g. serum). Antibodies specific to the coated antigen bind and non-specific antibodies are washed off of the surface. Biotinylated secondary antibodies recognize the Fc region of analyte antibodies, and horseradish peroxidase (HRP)-conjugated streptavidin binds to the biotinylated secondary antibodies. Alternatively, secondary antibodies can be directly conjugated to HRP. When an HRP substrate is added to the wells, the substrate reacts and produces a colorimetric signal that is proportional to the amount of analyte antibody bound to antigen in the well.

Multiplexed ELISA platforms employ a printed array of antigens (ELISA-array) (8, 9, 13, 14, 24–26), microspheres coated with antigens or antibodies (bead-based ELISA) (27–30), or a cocktail of antigens (cocktail ELISA) (19, 31–33). ELISA-array and bead-based ELISA platforms spatially separate the antigens on a surface, whereas conventional and cocktail ELISA coat the whole surface with the same antigen(s). Multiplexing with ELISA-array or bead-based ELISA offers higher specificity and sensitivity over a cocktail ELISA. Sensitivity of the cocktail ELISA falls with an increase in the number of antigens in a cocktail, which may be due to cocktail antigens blocking each other from binding to the plate (14). Commercial multiplex serology platforms include microsphere-based platforms Luminex and the substrate-printed ELISA-array platforms by RayBiotech and Scienion.

There is an outstanding need to deploy multiplex serology platforms across the world, including in low- and medium-income countries. The commercial platforms require specialized equipment for reading the assay and analyzing the output. The analysis software is closed source and difficult to extend and automate. Open, extensible, and low-cost multiplex serology platforms can enable widespread deployment and refinement of epidemiological, longitudinal, and diagnostic serological studies.

Here we report multiSero - a multiplex ELISA platform consisting of piezo-printed ELISA-arrays (8), a plate reader, and an open-source analysis pipeline, pysero. The ELISA-array can be read using an existing plate reader, any microscope with a motorized stage, or an open plate reader (Nautilus) that we report, ergo the platform is functional with equipment accessible in many laboratories once the ELISA-arrays are printed. Pysero accepts a variety of image file formats and is agnostic to the plate reader used for data acquisition and performs better than commercial analytical platforms for printed ELISA arrays. Pysero also enables visualization and interpretation of antibody responses. The data we report were acquired on Scienion plate reader and Nautilus, a plate reader designed with low-cost components to increase accessibility to this technology (34). Thus, this platform is usable by a large range of research and public health studies.

multiSero was validated by comparing total antibody titers in 114 sera from COVID-19 RT-PCR positive individuals and 87 COVID-19 negative sera banked before the SARS-CoV-2 pandemic. We report sensitivity and specificity of classification with each antigen, with Spike and receptor binding domain (RBD) of Spike performing with the highest sensitivity, consistent with earlier reports (35–37). We report a normalization method for comparing titers across plates to eliminate the need for a standard curve on each multiplexed ELISA plate. We trained classifiers using responses to multiple antigens and found that Spike protein suffices to classify COVID-19 positive and negative sera in our cohort. Our results demonstrate a performant and accessible multiplexed ELISA analysis pipeline ready for multiple applications, including serosurveillance, identification of viral proteins that elicit antibody responses, differential diagnosis of circulating pathogens, and immune responses to vaccines.

## Results

### A: Overview and components of multiSero

In our assay, plasma samples are added to a plate in which each well contained an array of antigens instead of a single antigen (fig. 1A). A panel of SARS-CoV-2 antigens was chosen to develop multiSero. The ELISA-array was printed using a Scienion sciFLEXARRAYER S12 (fig. 1B). The ELISA assay similar to conventional indirect ELISA was performed to enable colorimetric readout of antibody concentration. The assay is read out using a plate reader, either the SciREADER CL2 (Scienion) or Nautilus (fig. 1C; fig. S1). The plate reader used for data reported in each figure is noted in the caption. The images of ELISA-arrays were analyzed using pysero (fig. 1D) or the analysis package included with SciREADER CL2. In addition to antigen spots, ELISA array includes positive control, negative control, and fiducial markers in each well.

**Fig. 1.**
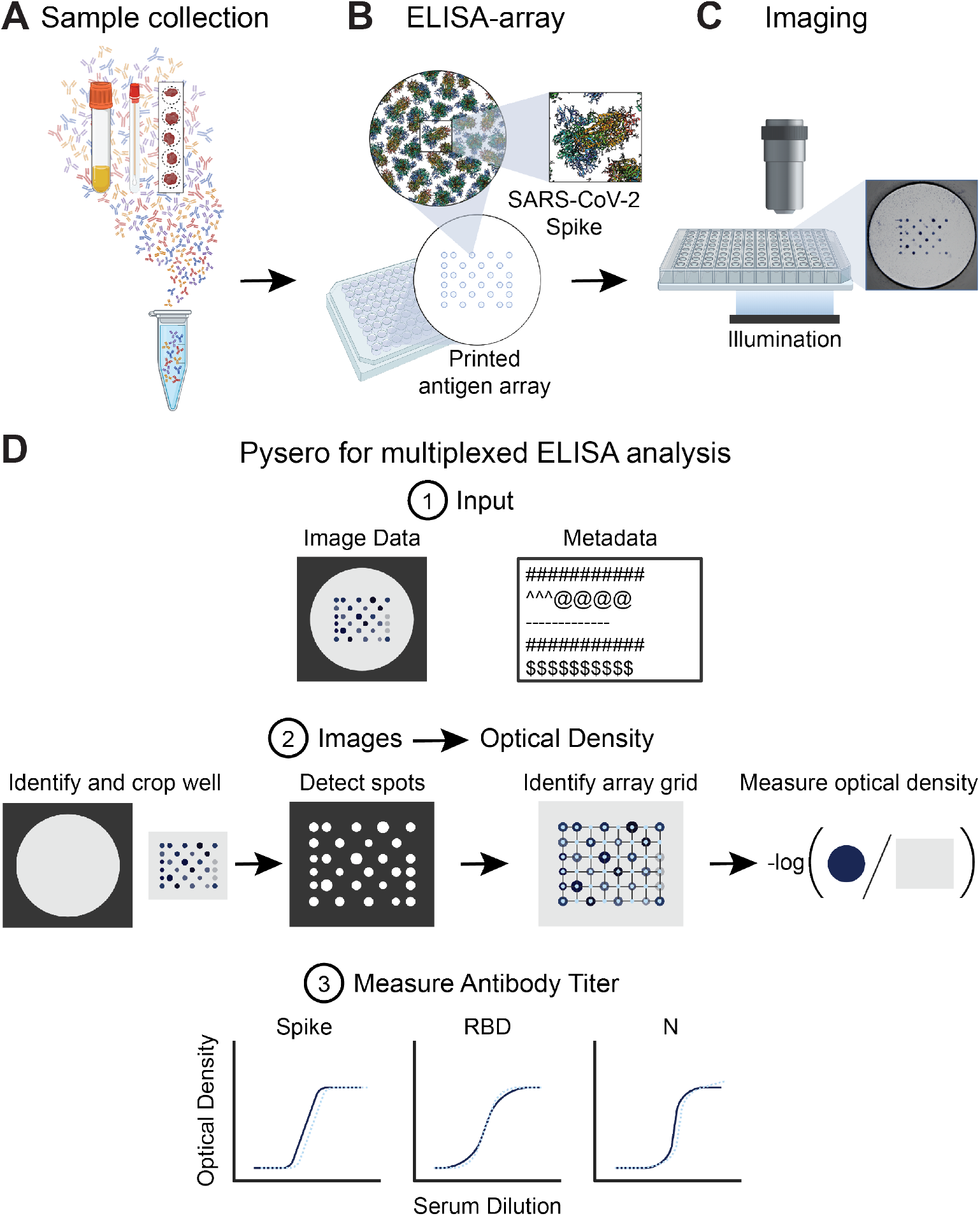
Overview of multiSero pipeline: **(A)** Sample containing antibodies derived from serum, saliva swabs, dried blood spots, or other source are overlaid onto antigen array **(B)** printed by a protein arrayer such as Scienion sciFLEXARRAYER S12. Each spot in the array contains a concentration of a single protein or protein domain. **(C)** The ELISA is performed and then the array is imaged using Nautilus plate reader or alternative reader. **(D)** Pysero is used to analyze printed substrate multi-antigen ELISA-arrays. The software takes well images and a metadata file describing experimental conditions and imaging parameters as input (D1). The images are auto-cropped around the antigen array. Antigen spots are detected and a grid is registered to the spots. Optical densities are computed from the spots that align with the registered grid. Optical density is computed as the -log of the ratio of spot intensity to the background intensity (D2). Sample antibody titer against each antigen in the array can then be measured based on the ODs of the controls in the assay (D3).

The Nautilus plate reader is an adaptation of the Squid microscopy platform with long-range XY stage that enables automated imaging of 96-well plates (fig. S1). The Squid platform is a performant, extensible, and open alternative with high-end microscopes with on-board computation. Pysero is a performant, extensible, and open-source alternative to commercial ELISA analysis software. Pysero provides modules that estimate antibody titers (fig. 1D, step 1) from images of the wells and metadata, as well as modules for interpretation of antibody titers. These modules are combined into pipelines for automated analysis of experiments. Nautilus and pysero together provide a complete solution for reading and analyzing developed ELISA-arrays.

As reported later, Pysero performs as well as the commercial analysis software and is independent of the plate reader (fig. 2). In addition, pysero provides machine learning classifiers that combine measurements from multiple antigens and multiple experiments. Data acquired with Nautilus is as informative as the commercial plate reader (fig. S6), the Scienion SciREADER CL2. Nautilus costs less than $1500, is portable, and can be built with off-the-shelf parts. The bill of materials is provided in table S1 and the plate reader is described in the Materials and Methods.

**Fig. 2.**
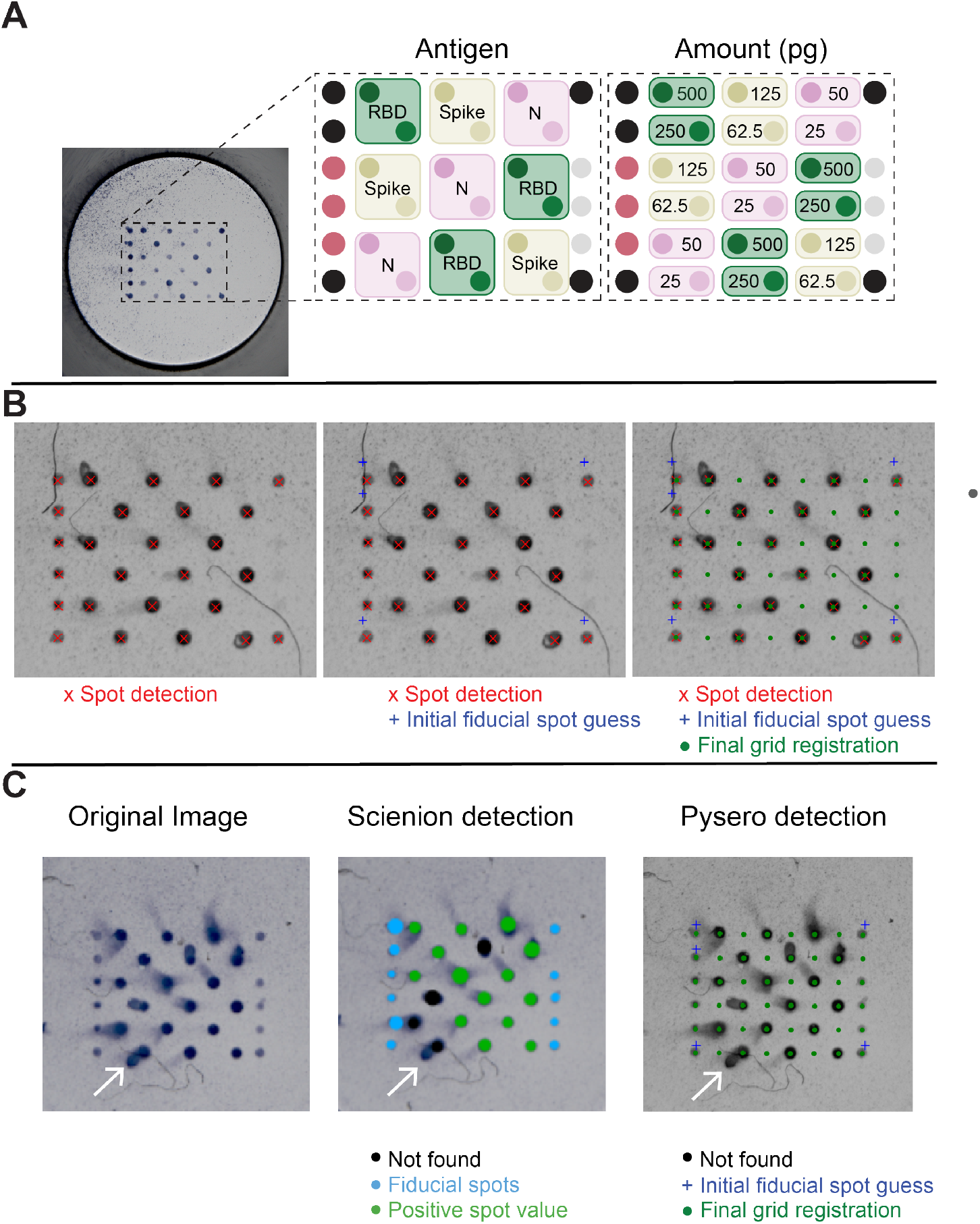
ELISA-array for SARS-CoV-2 and its analysis with pysero. **(A)** Image of well containing Scienion-printed antigen array. Array layout shows the relative locations of SARS-CoV-2 antigens included in the array: receptor binding domain of spike-RBD (RBD; green), spike (ochre), and nucleoprotein (N; pink). Each antigen was spotted at two concentrations which are indicated in the right panel of the layout. Black spots represent anti-kappa-biotin fiducials, mauve spots represent anti-IgG Fc, and grey spots represent GFP foldon. **(B)** Pysero first detects center points of all spots in the cropped image (red x). Fiducial positions (blue +) are initialized based on detected spot coordinates. Coordinate transformation that registers the initial fiducials with detected fiducials is estimated using particle filtering. A grid containing all spot locations (green) in the antigen layout is then transformed onto the image using the estimated coordinate transformation. The coordinates of each spot are then used to crop individual spots and extract their OD. **(C)** Comparison of Scienion analysis and pysero spot detection. Original image of a well imaged by SciREADER CL2 includes comets (white arrow) and fiber-like debris. Center image is the output of Scienion spot detection and analysis. A black spot indicates an analyte-containing spot was not found at the location, green spots indicate positive spot value, and blue spots indicate the location of fiducial spots. The size of the spot indicates the area analyzed by Scienion. Right image is the output of pysero. Green dots indicate registered grid positions (size of marker unrelated to area analyzed), blue crosses indicate initialized fiducial spots.

We summarize key components of the pysero here and describe them in detail in Materials and Methods. The analysis module (fig. 1D, step 2) estimates antibody titers from the images and ELISA-array metadata. In this step, the antigen array is cropped, spots are detected, and each spot is assigned an antigen identity based on ELISA-array metadata. The detection of spots and assignment of identity rely on a grid registration algorithm that uses the fiducial spots as landmarks. The fiducial 2 spots are deposits of antibodies whose signal intensity is invariant in the assay. Individual antigen spots are then cropped, the intensityspot 3 background around each spot is estimated, and the optical density (OD) is computed as 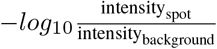. The interpretation module takes OD and experimental metadata as inputs and enables measurement and visualization of antibody concentration.Commonly used OD vs. dilution plots (fig. 1D, step 3) provide an estimate of relative antibody titers for each antigen in each sample. The absolute titer (ng mL^−1^) of antibodies can be semi-quantitatively determined by running a standard of known concentration on the plate and interpolating OD of corresponding antigen into the standard curve. The interpretation module also includes machine learning-based classifiers that pool the ODs from multiple antigens and multiple experiments to classify a given sample as COVID-19 positive or negative.

### B: SARS-CoV-2 ELISA-array and its analysis by pysero

We included Spike, RBD, and Nucleoprotein (N) at two concentrations (fig. 2A) in our SARS-CoV-2 antigen array as detailed in Materials and Methods. These antigen concentrations were chosen to ensure that the antigen was present in excess of the antibodies in the sera and the dynamic range of antibody amounts led to a measurable dynamic range of OD. Future refinement of the assay would include a single optimized concentration per antigen. In addition to the SARS-CoV-2 antigens, fiducial spots consisting of biotinylated anti-kappa light chain antibodies were added to the array to facilitate grid registration and subsequent analysis. The stepwise process of grid registration using fiducials is shown in fig. 2B. The fiducial spots were printed at a constant concentration and the streptavidin-HRP recognized the fiducial spots uniformly. Anti-IgG antibody was spotted as a positive control to recognize total IgG in serum, and GFP foldon was used as a negative control, previously described by Waltari et al. (8) Introduction of artifacts is possible while performing multiplexed ELISA, and common artifacts include comets and debris. Both comets and debris are present in the representative image read by a SciREADER CL2 in fig. 2C. The layout and spacing of the antigen array was such that experimental artifacts, such as smearing of spots (comets, example in fig. 2C), would minimally affect the analysis of adjacent spots.

We compared the performance of the SciREADER CL2 software and pysero in recognizing the spots in the array. The SciREADER CL2 failed to identify four spots that were partially obfuscated by comets, whereas pysero detected these spots (fig. 2C). Furthermore, we compared spot intensities, background intensities, and ODs derived from the SciREADER CL2 software and pysero, and found similar trends (fig. S2). Pysero was more robust to comets relative to the SciREADER CL2 software in two respects: spot detection and background estimation. The SciREADER CL2 software detected spots that were larger than the central spot when the spot had a comet which resulted in the inclusion of background pixels in the spot region. Including background pixels in the spot region results in higher intensity, and thus lower OD. The SciREADER CL2 background estimation was also affected by comets in that it estimated background from the pixels surrounding detected spots assuming circular spot shape. Therefore, the SciREADER CL2 analysis platform underestimated the background intensity when there was a comet, and this resulted in lower OD. Additionally, the Scienion analysis platform is only capable of analyzing images acquired using the SciREADER CL2, prohibiting a comparison of Nautilus images and SciREADER CL2 images analyzed by the SciREADER CL2 software.

We evaluated whether the comets bias the ODs measured with pysero by comparing the duplicate spots, i.e., spots from the wells that contained the same antigens and same serum (fig. S3). We find that the presence of comets does not cause observable bias or variance in ODs measured with pysero. In sum, pysero enables more robust analysis of antibody titers than the commercial analysis software available with SciREADER CL2.

### C: Limits of detection and quantitative analysis

Each antigen on the array has an independent lower limit of detection which is influenced by the amount of non-specific serum protein binding to each antigen in the sample. We determined the lower limit of antibody detection for each antigen in human serum by evaluating sera from two cohorts: a group of individuals from the Long-term Impact of Infection with Novel Coronavirus ([LIINC](https://www.liincstudy.org/)) cohort, all of which tested positive for SARS-CoV-2 by qRT-PCR test, and sera acquired from a blood bank that was banked before SARS-CoV-2 was circulating in the population. We termed the serum pool from the LIINC cohort as “Positive Pool”, and the serum pool from the pre-pandemic sources as “Negative Pool”.

We first evaluated which of the two amounts of Spike, RBD, and N proteins are suitable for detection of antibody titers in SARS-CoV-2 positive and negative sera in our cohort. ELISA with dilution series of the Positive Pool and the Negative Pool (fig. S4) showed that 50 pg of N (nL spotted at 50 µg mL^−1^), 250 pg of RBD (1 nL spotted at 250 µg mL^−1^), and 62.5 pg (1 nL spotted at 62.5 µg mL^−1^) of Spike transition from not being detectable at low concentrations of Positive Pool to being saturated at high concentrations of the Positive Pool while these amounts of antigen remain unsaturated by the Negative Pool at all serum concentrations. This data clarified that using the lower of the two amounts we spotted for each antigen suffices to discriminate antibody titers between positive and negative sera and clarified appropriate serum dilutions for accurate antibody titering given the amount of antigen spotted.

Results for antibody responses against 250 pg of spotted RBD are shown in fig. 3A-D and all other antigens are summarized in table S2. To determine the relative range of antibody responses to each antigen, multiplexed ELISA was performed using a dilution series of the Positive Pool and Negative Pool (fig. 3B). For a given serum dilution, a sample is positive for antibodies against a SARS-CoV-2 antigen if the unknown sample’s OD is three standard deviations above the mean Negative Pool OD at that dilution.

**Fig. 3.**
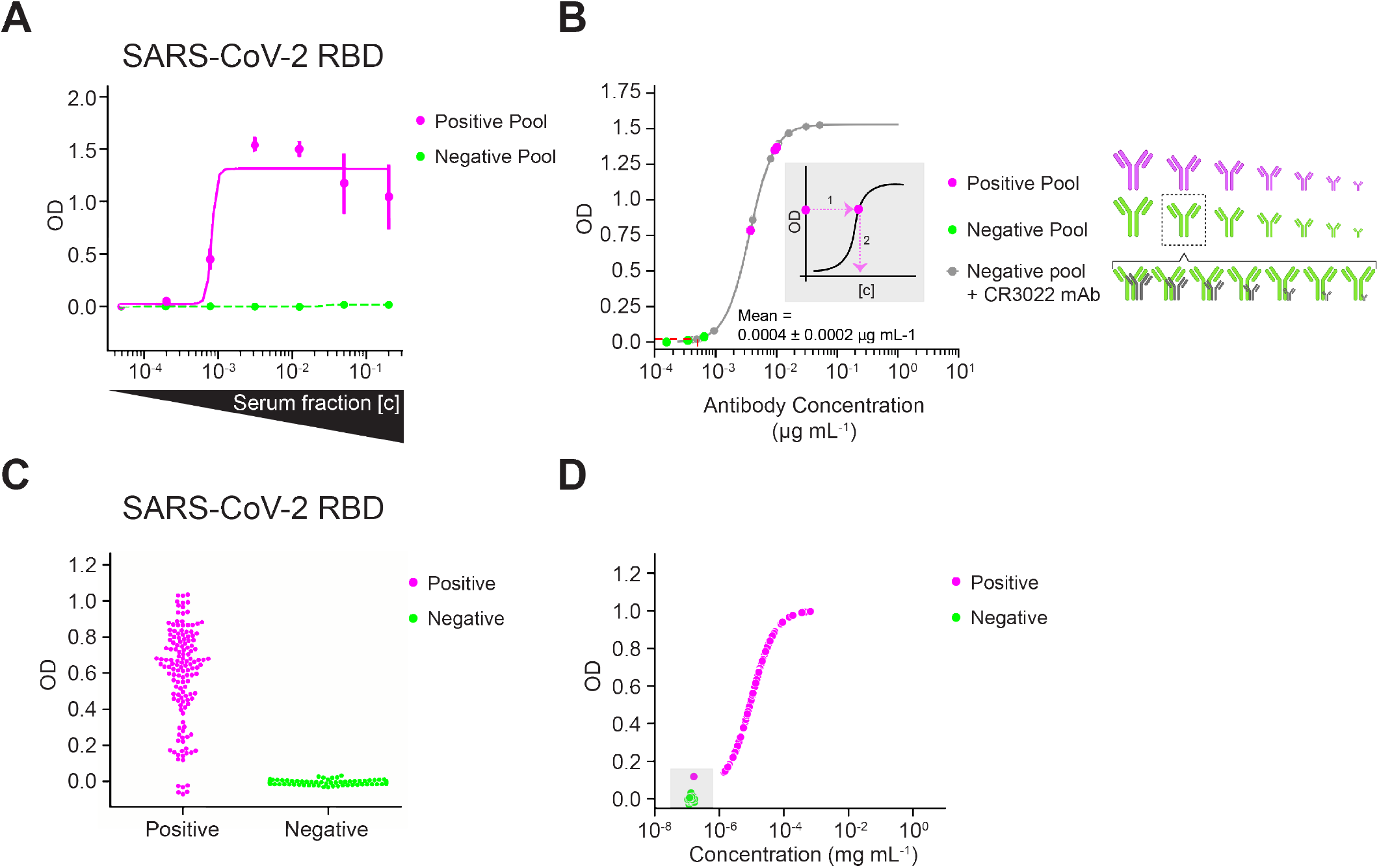
Determining limits of detection in the presence of complex background and validation by SARS-CoV-2 epidemiology survey. **(A)** ODs denoting antibody response against RBD antigen spot (250 pg). Serial dilution of sera pooled from RT-PCR-positive cohort (magenta solid line) and a pooled RT-PCR-negative cohort (green dashed line). **(B)** Concentration of antibodies against RBD spotted at 250 pg present in pooled positive sera (magenta points) and pooled negative sera (green points) at 1/200 dilution. Grey curve and points denote standard curve (OD vs known antibody concentration) obtained from serial dilution of mAb CR3022 in the 1/200 diluted Negative Pool. The antibody schematic at right illustrates 7-point serial dilutions of the pooled positive sera (magenta), the pooled negative sera (green), and the mAb CR3022 (grey) in the 1/200 diluted Negative Pool. The ODs corresponding to the Positive Pool and Negative Pool were interpolated into the standard curve, and the resulting antibody concentration of the unknowns can be read out from the x-axis using the fit of the standard curve. The mean antibody concentration of the Negative Pool (1/200 serum dilution) is indicated on the plot (mean = 0.0004 *±* 0.0002 µg mL*−1*; red dashed line), and the lower limit of detection for a positive result is 3 standard deviations above this mean. Data for panels (A-B) were acquired using SciREADER CL2. **(C)** Range of ODs of RT-PCR-positive individuals (magenta) and RT-PCR-negative individuals (green) from a small episurvey using samples from the LIINC cohort. Each point represents one triplicate for each antigen. **(D)** ODs from individual serum samples interpolated into mAb CR3022 standard curve. Sera are colored according to their RT-PCR-positive or RT-PCR-negative status. An OD that is outside of the range of ODs corresponding to the standard curve cannot be quantified. Points that are lower than the lowest mAb CR3022 concentration are plotted by OD and arbitrarily at 10^−7^ mg mL^−1^. Data for panels (C-D) were acquired with Nautilus.

Quantitative measurement of antibody titer to each antigen in the array varies based on antibody affinity and concentration. To estimate the antibody titers specific to RBD and Spike, we generated a standard curve with a monoclonal antibody (mAb) CR3022 (38), which recognizes a conserved epitope of RBD of SARS-CoV-2 and SARS-CoV. The CR3022 titration was performed using the Negative Pool as a diluent to match the matrix found in human sera. Figure 3B shows OD vs serial dilutions of the mAb CR3022 in the range 1 − 0.00024 µg mL^−1^ (fig. 3B; grey points). We also show parameter logistic regression of this data (fig. 3B; grey points), which allows us to interpolate the reactivity of antibodies in sera from the measured ODs (fig. 3B; inset). For 250 pg of spotted RBD, the data indicates: a) the lowest concentration of CR3022 in Negative Pool (0.2ng mL^−1^) has reactivity equivalent to the Negative Pool alone (fig. 3B), and b) the Negative Pool on average has reactivity equivalent to 0.4 ng mL^−1^ of CR3022. Antibody responses against other antigens are shown in table S2. Using this assay, we can qualitatively determine whether individual samples contain antibodies against specific antigens and also approximate the amount of sample reactivity by equating OD to the equivalent of mAb CR3022 recognizing RBD at a known concentration and binding affinity.

Multiplexed ELISA was performed on the individual serum samples to determine the range of antibody responses against SARS-CoV-2 (fig. 3D). We set the positivity threshold for specific antibodies against RBD in this assay at 1.1 ng mL^−1^, i.e., three times the standard deviation of the mean binding reactivity of the Negative Pool. Three samples from individuals with positive qRT-PCR results had lower antibody concentrations against RBD than the lowest concentration on the mAb standard curve, 1.6 ng mL^−1^ (fig. 3D; grey inset). These samples had low responses to all antigens in the array, suggesting these individuals had sub-optimal antibody responses to this infection.

### Normalization methods for large-scale multiplexed ELISA data

One approach to quantitative analysis of reactivity is to incorporate a set of standard antibodies as we reported. But, incorporating a set of standards to generate a standard curve per antigen in the array each time multiplexed ELISA is performed is reagent-heavy and labor-intensive. The use of standard curves also reduces the number of samples that can be run per plate. An attractive alternative is to normalize the ODs of each experiment to the experiment that included the standards. ELISA array provides fiducials and positive controls that facilitate such data normalization. Proper normalization is also necessary for pooling data from across the experiments for statistical analysis, e.g., training more accurate diagnostic classifiers.

We explored normalization of multiplexed ELISA OD values to enable pooling of data across plates without the need to run a standard curve per plate. We explored 3 normalization methods: The ODs of spots in duplicate plates were normalized with the ODs of fiducial spots (biotinylated anti-kappa light chain antibodies that are detected by the streptavidin-conjugated secondary antibody) per plate, the ODs of the anti-IgG Fc spots per plate, or the ODs of anti-IgG Fc spots per well (fig. S5). Each spot location in the array was compared to the spot with the same array location in a duplicate plate. Without normalization, duplicate plate datasets exhibited low bias and noise. After normalizing by the anti-kappa light chain fiducial signal present in each plate, bias and noise were marginally reduced for one out of three datasets, with bias = -0.074 and noise = 0.108 without normalization compared to bias = -0.023 and noise = 0.082 after normalization. Normalizing by anti-IgG Fc signal present in each plate resulted in a subtle reduction in bias and noise consistently in all 3 datasets. Normalizing the OD values in each well by the anti-IgG Fc signal present in the same well decreased the bias in 2 out of 3 datasets but considerably increased the noise in each dataset. The best normalization method provided only a small improvement over no normalization. These experiments were performed using the same reagent lots and in a short span of time. ELISA-array data accumulated over weeks or months and using multiple lots of reagents are expected to have larger variability and can benefit from these normalization procedures.

### D: Sensitivity and specificity of SARS-CoV-2 multi-antigen ELISA

To determine the best antigen or antigen set for discriminating positive and negative sera, we trained gradient boosting tree classifiers using RT-PCR status of the sera as the ground truth. We used data from 189 sera (106 positive, 83 negative). All sera were assayed at least in duplicate, some of them in quadruplicate and were spread over 6 different plates. The robustness of trained classifiers was evaluated using receiver operating characteristic (ROC) curve and the area under the curve (AUC). Sensitivity of the assay at 95% and 99% specificity using single antigens are reported for each antigen in table S2 (N=507). In fig. and fig. S6, we report ROC curves and sensitivity at 95% specificity.

We evaluated if the choice of plate reader affects the performance of the assay. We used pysero to compare sensitivity and specificity of classification of the COVID-19 positivity of samples by analyzing the data acquired with SciREADER CL2 and Nautilus (fig. S6).The equivalent classification performance illustrates that the data acquired with Nautilus is as informative as commercial solutions and pysero can be used with any plate reader.

To test if incorporating information from multiple antigens improves the classification performance, we trained a gradient boosting tree classifier on 60% of the SARS-CoV-2 positive and negative serum data (training and validation sets) in the cohort and performed classification on the remaining 40% of the dataset (test set). The ROC curves for each antigen and the classifier were generated on the test set (N=189) (fig. 4). The ROC curve reports the true positive rate (sensitivity) and false positive rate (1 - specificity) as a function of the OD threshold (for single antigens) or probability threshold (for the classifier) used to classify the serum samples. RBD and spike show similar performance (97.7-97.8% sensitivity) for both spotted concentrations. Nucleoprotein shows worse performance (95.5% sensitivity at 95% specificity) (fig. 4A). Using this analysis, users can select OD thresholds depending on the metric (i.e. sensitivity or specificity) more appropriate for their application. For example, first-pass surveys to screen for potential positives in a large cohort may tolerate a higher probability for false alarm to capture all positives in the initial screen in lieu of the highest possible sensitivity. Figure 4B illustrates how true positive rate and false positive rate change as the threshold value varies.

**Fig. 4.**
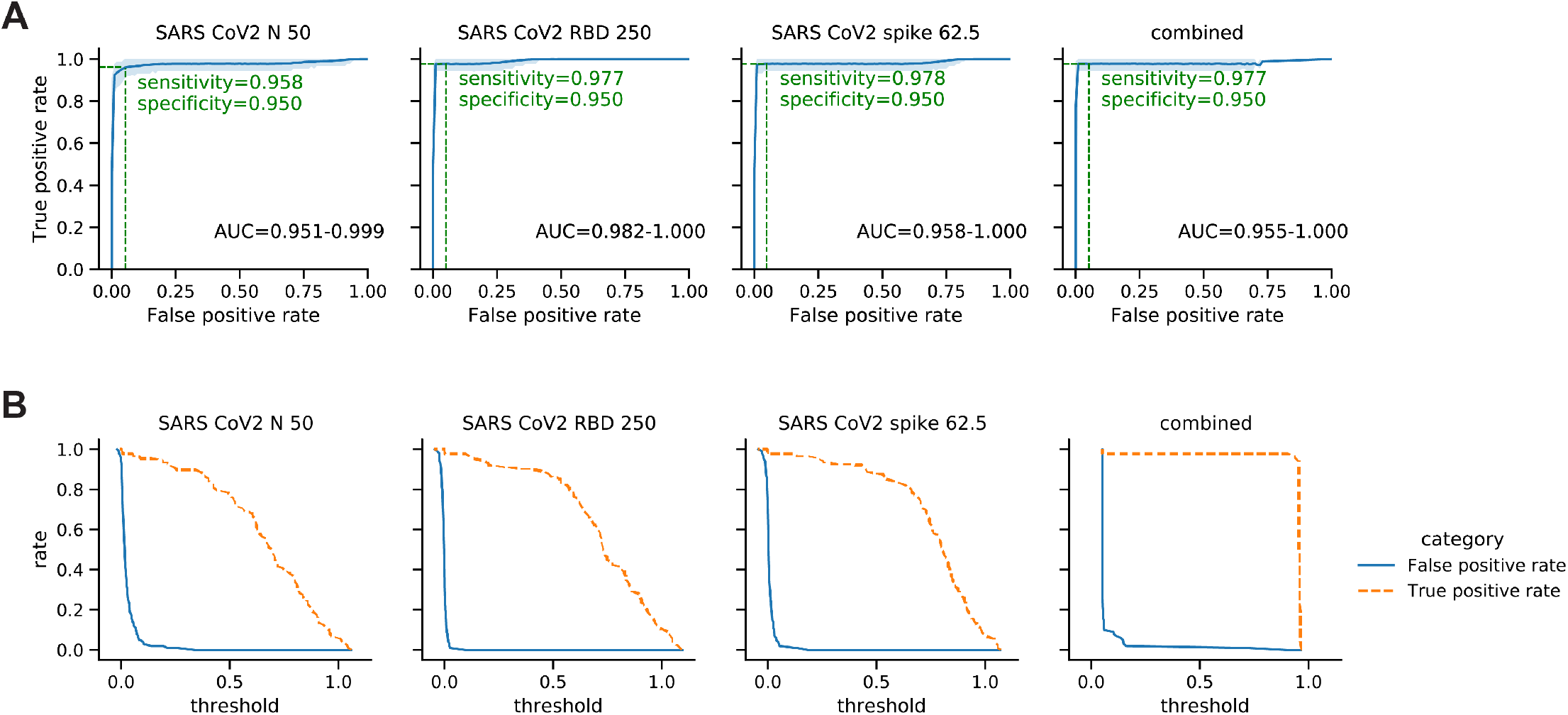
Performance of classifiers that use single and multiple antigens to classify positive and negative sera. **(A)** ROC curves (blue line) for classifiers that use single antigens (left to right: nucleoprotein, RBD, and spike) and a classifier that combines information from multiple antigens. Blue shades represent 95% confidence intervals of ROC curves. For each antigen, the sensitivity (True positive rate (TPR)) at 95% specificity (1 - false positive rate (FPR)) is denoted (green dashed line) on the curve. 95% confidence interval of area under the ROC curve (AUC) is reported below each curve. **(B)** TPR (orange dashed line) and FPR (blue line) represented as a function of the OD threshold (for single antigens) or probability threshold (for the classifier) used to classify positive and negative sera. ODs were normalized by mean anti-IgG Fc signal per plate. 189 sera (106 positive, 83 negative) were assayed at least in duplicate, some of them in quadruplicate, spread over 6 different plates. Data for fig. 4 were acquired with Nautilus.

The classifier that uses ODs from all antigens performed with 97.7% sensitivity at 95% specificity, which is comparable to the best performing single antigens (RBD and Spike). Examining the images and ODs of the false negative classifications showed low OD across all antigens in both replicates, which clarifies why incorporating information from all antigens did not help correct these false negative samples. On the other hand, all false positive serum samples showed relatively high OD values for some antigens but only in one of the duplicates, indicating these false-positive samples most likely result from high background that randomly occurs during the assay, and could be eliminated by including more replicates.

## Discussion

multiSero platform provides commercial-grade sensitivity and specificity with an in-house surface-printed antigen array, open source analysis software (pysero), and open source plate reader (Nautilus). Our multiplexed ELISA assays of SARS-CoV-2 antigens illustrate that this open pipeline provides performance comparable to commercially available platforms and provides features that are not available in commercial platforms. Commercially available printed-substrate ELISA-arrays either require a proprietary plate reader and analysis software or they are compatible with laser scanners and software used for protein microarrays (Raybiotech, Atlanta, GA; Quansys, Logan, UT; Scienion, Berlin, Germany; Abcam, Cambridge, MA; Meso Scale Diagnostics, Rockville, Maryland). Because pysero is agnostic to the image source, it can also be used to analyze data from a variety of surface-printed array products including those that use fluorescence as readouts. Here we have tested pysero with data acquired using Scienion’s SciREADER CL2 and the Nautilus plate reader.

The described SARS-CoV-2 multi-antigen array is useful as an epidemiology survey tool as it has increased throughput compared to conventional ELISA by increasing the number of analytes per plate. The in-well biotinylated anti-kappa light chain spots, GFP foldon spots and anti-IgG Fc spots function as internal controls and decrease the sample volume needed per assay. The sample size of this study is small; however, sensitivity at 100% specificity will increase as this array is more routinely used for epidemiological surveillance.

Accuracy of analyzing the surface-printed array can be improved through minimization of artifacts such as comets. Comets are smearing of spotted antigen:antibody complexes and are true signal (fig. S3), so it is important to integrate the signal at the registered center of the spot and the comet for the most quantitative result. Currently pysero only extracts OD surrounding the registered center of the spot. Comets are common to surface-printed protein arrays, and one hypothesis is that comets form when the surface is not completely dry before performing the ELISA. In our hands, waiting up to 30 minutes for the plate to equilibrate at room temperature before the experiment modestly reduced comets, suggesting alternative sources of comets. In anticipation of comets, we implemented 2 practical solutions to reduce effects of comets regardless of their source. First, we designed an antigen array layout in a checkerboard pattern so that comets would not overlap with nearby spots and interfere with OD extraction. Second, each antigen is in triplicate within the array and the layout is such that each triplicate location is distant from the other triplicates. Future iterations of this assay may explore alternative array technology to ameliorate comet troubleshooting. For example, the surface of the plate, plate porosity, or the formulation of the protein stock for printing could be altered to prevent mobility of the spot, or microfluidic devices could be used to control flow of sera and reagents as well as isolate antigen reservoirs.

Multiplexed ELISA and open-source software have high potential for surveying exposure to multiple viruses in low-resource regions with endemic infectious disease. Many infectious diseases (not only viral) manifest with nonexclusive symptoms, and a multiplexed ELISA-array with antigens representing circulating pathogens can be used to simultaneously evaluate the prevalence of multiple pathogens. The approach of using surface-printed protein arrays and open source analysis tools can be adapted for multiplexed detection of pathogens by printing pathogen-specific antibodies, instead of antigens.

Non-contact arrayers can be purchased for $100,000-300,000. The printing technology is the most expensive aspect of the pipeline, and access to the antigen array printer is a limiting factor in wide-spread deployment of multiplexed ELISA to resource-poor regions. Advances in bioprinting technology enable the possibility of an open arrayer (39). Partnerships with non-governmental organizations and multi- or bi-lateral agencies with global health missions can provide support for hardware or cold chain storage of surface-printed arrays. Alternatively, arrays with more intrinsic stability and portability can be devised. Two such examples are the origami-inspired foldable paper protein arrays or microfluidic devices (40–42), which would also rid the current platform of analysis challenges due to comets.

A combined classifier was trained on responses to all array antigens using sera from confirmed SARS-CoV-2 positive individuals and negative sera banked prior to the COVID-19 pandemic. We chose the gradient-boosting framework since it enables interpretation of feature importance. We find that there is no advantage in classifying the positive/negative cases when responses to multiple antigens are combined, because the single best antigen, Spike spotted at 62.5 pg, provided high discriminatory power. The combined classifier may increase the specificity and sensitivity for multiplex assays that have less specific responses. For example, less expensive serology pipelines, such as ReScan (11) employ linear peptides that interact with lower specificity than the folded proteins. Our approach of building an interpretable classifier may improve the specificity and sensitivity of such assays. To our knowledge, other analysis programs for multi-antigen ELISA currently do not offer combined analysis of multiple antigens in order to make positive/negative calls of a sample.

In summary, multiSero provides a performant and scalable pipeline consisting of open tools that can enable multiple diagnostic and epidemiological studies: serosurveillance, differential diagnosis of circulating pathogens, detection of viral proteins, and immune responses to vaccines.

## Materials and Methods

### A: Recombinant protein production

Plasmids encoding SARS-CoV-2 Spike (S) ectodomain or Receptor Binding Domain (RBD)36,41 were transiently transfected into suspension Expi293 cells at 1-1.5 L scale in 2.8 L Optimum Growth Flasks (Thomson Scientific), following the manufacturer’s guidelines. Three days after transfection, cell cultures at >75% viability were centrifuged at 500 g for 30 min, followed by filtration of the supernatant using a 0.45 um NalGene Rapid Flow filter unit. The supernatant was adjusted to pH 7.4, and directly loaded onto a 5 mL HisTrap Excel column pre-equilibrated with 20 mM sodium phosphate, 500 mM NaCl, pH 7.4 using an AKTA Pure. Captured proteins were washed with 60 column volume (CV) of (20 mM sodium phosphate, 500 mM NaCl, 20 mM imidazole, pH 7.4) and eluted with 10 CV of (20 mM sodium phosphate, 500 mM NaCl, 500 mM imidazole, pH 7.4). Eluted proteins were buffer exchanged into PBS using either 3kDa MWCO (for RBD) or 100 kDa MWCO (for Spike) Amicon concentrators, supplemented with 10% glycerol, and filtered prior to storage at -80°C. Protein stability after freeze thaw was confirmed by analytical SEC-MALS.

A codon-optimized His tagged SARS-CoV-2 Nucleocapsid (N) gene (43) was synthesized and cloned into a pET-28 vector by Twist Bioscience (San Francisco, CA, USA). The expression plasmid was transformed into T7Express bacterial expression cells (New England Biolabs, Ipswich, MA) and a single colony was used to grow 10 mL of overnight culture in LB/Kanamycin, which was then used to inoculate 1 L of LB/Kanamycin. The 1 L culture was shaken at 37°C, 200 rpm until the O. D. 600 reached 0.6. The temperature of the culture and incubator were lowered to 25°C and protein expression was induced by addition of 0.5 mM Isopropyl-β-D- thiogalactoside (IPTG). After 20 h, the bacterial cells were isolated by centrifugation and the pellet resuspended with Buffer 1 (50 mM phosphate, pH 8.0, 1 M NaCl, 10% glycerol). The cells were lysed by sonication after a 0.5 h treatment with lysozyme (MilliporeSigma, Burlington, MA), benzonase nuclease (MilliporeSigma), and 1 cOmplete EDTA-free Protease Inhibitor Cocktail (MilliporeSigma). The clarified lysate was loaded onto a 1 mL HisTrap Fast Flow column (Cytiva, Marlborough, MA) at a flow rate of mL/min. The column was washed with 100 column volumes (CV) of Buffer 1 + 50 mM imidazole and pure protein eluted in 20 CV Buffer 1 + 250 mM imidazole. Fractions containing pure protein were buffer exchanged using PD-10 desalting columns (Cytiva) into storage buffer (50 mM phosphate pH 8.0, 500 mM NaCl, 10% glycerol), aliquoted, and frozen at -80°C.

### B: Printing of protein arrays

All protein arrays were printed using a Scienion sciFLEXARRAYER S12 instrument, following general instrument parameters described previously (8). Proteins were aspirated from a 384-well source plate (Scienion) maintained at dew point, and dispensed onto Fluotrac™ 600, Greiner Bio-One 96-well plates (Fisher Scientific, Waltham, MA) maintained at 60% humidity and ambient temperature, with drop volumes of 330-350 pl and 2 drops per spot. Coronavirus Spike, RBD and Nucleocapsid proteins were printed in either a 6×6 grid or 8×6 “checkerboard” pattern. Additional biotinylated anti-kappa light chain spots, GFP foldon spots, and anti-IgG Fc spots were also printed as fiducials, negative controls, and positive controls, respectively. The printed arrays were maintained at 75% relative humidity and ambient room temperature overnight to allow for adsorption of the proteins to the surface of the plate, followed by vacuum sealing and storage at 4° C until use.

### C: Open plate reader, Nautilus

The plate reader, nick-named Nautilus, was derived from the open microscopy platform [squid](https://squid-imaging.org/) (34). The scanning unit was adapted to hold 96-well plates and an LED array, diffuser, and 650 nm filter were added to evenly illuminate the wells. The imaging setup consists of a 4x/0.1NA objective (Boli Optics), an f = 35 mm imaging lens (Hivision) as tube lens and an industrial camera with monochrome Sony IMX226 CMOS sensor (Daheng). While motorized autofocus has been implemented, the data is collected with an early version of the system where manual focus is performed for well A1 before automated scanning of the entire plate starts. Images are saved as 8-bit bmp files for downstream processing. The system is controlled with a python-based graphical user interface, which is part of the squid framework and can be further integrated with the pysero processing pipeline.

### D: Multiplex ELISA analysis with pysero

The source code for pysero is available at https://github.com/czbiohub/pysero

#### Detection of spots

Given a printed microarray with known fiducial and sample grid coordinate locations, the pysero spot detection pipeline will extract those locations from the metadata.xlsx file as well as other critical parameters such as number of rows, columns, vertical and horizontal pitch, spot width and pixel size. Spot detection is a multi-stage process beginning with several preprocessing steps to assist detection when the sample can be dirty or noisy. First, pysero extracts the inner-well area from the image using a combination of multi-modal intensity thresholding such as “otsu” and “rosin” thresholds, then checking that the area meets minimum size and eccentricity constraints. From this cropped image, we apply a uniform 2D Laplacian of Gaussian filter to the whole image using a kernel size sigma that is a quarter the spot width in nanometers. Next, this filtered image is fed into the openCV module *SimpleBlobDetector* (44). The module generates a range of binarized images from thresholds based on minimum and maximum intensity, groups blob center coordinates across binary images, and we filter out relevant blobs using the *SimpleBlobDetector* parameters minimum circularity, convexity, distance between spots, and repeatability. The outputted coordinates of proposed spots are further filtered based on distance from the border of the image.

The final step is to align the detected spots to the known printed array configuration of fiducials as defined in the meta-data. Proposed spots may contain all spots in the array but may have missed some due to artifacts or lack of signal, and may contain false positives from noise. In order to find a robust fit between fiducial locations and proposed spots we use particle filtering (45), a sequential importance sampling method often used in tracking but can also be found in medical image segmentation applications (46). A particle is defined as a grid pose estimate, and we allow for translation, rotation, and scaling transformations when sampling particles. An initial grid estimate is based on the known physical size of the grid and is placed at the center of the image with 0 degrees of rotation and a scale of 1. A random set of N=4000 particles is generated and for each particle the sum of distances from fiducials to their nearest proposed spots is calculated. Weights are computed as the inverse of the summed distances and a new set of N particles is generated using importance sampling of these weights, with a small number of distortions added. This process allows for successful particles to proliferate while unsuccessful particles die out, and is repeated until convergence.

#### Measurement of OD

Once the fiducials of the array are identified using particle filtering, the expected positions of other spots can be determined by their positions on the grid. A simple threshold at 75% of pixel intensity level is performed within a bounding box around the expected spot position to segment the spot. The bounding box dimension is set to be twice as big as the expected spot intensityspot width to account for spot printing and fiducial fitting errors. If no spot is detected within the bounding box, a circular mask with the expected spot size and position is used to compute the spot intensity. To estimate the background of the image, the image is first divided into blocks of 128×128 pixels. The median of each block is computed, and a second order 2D polynomial function is fit to the blocked median to get the background image. The spot and background intensities are then defined by the median pixel intensity of the sample and background images within the segmented spot. The OD value of a spot is defined by 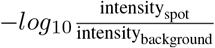. For computing ROC curves, each spot’s OD value is normalized by the average OD of anti-IgG Fc spots over the entire plate. OD values from the same antigen in the same well are then averaged. ROC curves and the area under the curve are computed using scikit-learn functions roc_curve and roc_auc_score. 95% confidence intervals of the ROC curves are computed by stratified bootstrapping the OD values, which are then used to generate distributions of ROC curves.

#### Classification model

The normalized OD values of each antigen, anti-IgG Fc, and GFP foldon spots in each well were used to construct the feature vector of each serum sample. Binary gradient boosting tree classifiers are built and trained using xgboost (47) to distinguish the positive and negative samples. 60% of serum data (N=318) were used to train the model. Model training and hyperparameters tuning were done by choosing the model with the best average validation AUC score from 4-fold cross-validation (75% training and 25% validation splits). The model performance was evaluated on the remaining 40% of the dataset (test set, N=189) that were not used for training and model selection.

### E: Validation of multiSero with COVID-19 sera

#### SARS-CoV-2 ELISA Array Assay

Plasma samples were assayed with the SARS-CoV-2 ELISA array as described in Waltari et al. (8) with modified parameters. Briefly, 200 uL blocking buffer (0.05% Tween, 0.5% bovine serum albumin fraction V, 2% filtered fetal bovine serum, 0.2% bovine gamma-globulin, 0.05% Proclin300TM, 5mM EDTA in PBS) was added to each well of the printed and cured ELISA array plates and allowed to incubate for 1 hour, then aspirated. 100 uL sample serum diluted in blocking buffer was added to each well and allowed to incubate 1 hour. Sample was aspirated and wells incubated for 1 hour with 100 uL of an even mixture of biotinylated goat antibodies targeting the kappa (25 ng mL^−1^, Southern Biotech cat. 2070-08) and lambda (25 ng mL^−1^ Southern Biotech cat. 2060-08) light chains. Wells were aspirated and incubated for one hour with 100 uL streptavidin-conjugated HRP (0.2 µg mL^−1^). Wells were aspirated and developed with 50 uL sciCOLOR T2 precipitating TMB reagent (Scienion). Wells were aspirated, sealed, and imaged shortly thereafter on a sciREADER CL2 plate reader (Scienion) or Nautilus plate reader. Wells were washed between steps with PBS 0.05% Tween.

#### SARS-CoV-2 positive samples and negative controls

The SARS-CoV-2 ELISA array assay was validated using plasma samples from RT-PCR confirmed SARS-CoV-2 infected patients from the Long-term Impact of Infection With Novel Coronavirus (COVID-19) ([LIINC](https://www.liincstudy.org/), NCT04362150) study. Various plasma samples collected before the COVID-19 pandemic were used as negative controls. All samples were stored at 4°C and diluted 1:1 in HEPES buffer (40% glycerol, 0.04% NaN3, 40 mM HEPES in PBS), and further diluted 100X in ELISA array blocking buffer before assays. Pooled SARS-CoV-2 positive samples and CR3022 monoclonal mouse antibody synthesized in-house (48) were serially diluted to titrate antibody concentrations in samples run on the same ELISA array plate.

## Data Availability

The images of SARS-CoV-2 ELISA-array assayed with 189 sera, spread over 6 plates, are available publicly via google drive (https://bit.ly/34ZjdtQ). The python code to analyze this data is available from this GitHub repository (https://github.com/czbiohub/pysero). The repository also includes code and instructions to reproduce fig. 4 from above images.

https://github.com/czbiohub/pysero

## Data Availability

The images of SARS-CoV-2 ELISA-array assayed with 189 sera, spread over 6 plates, are available publicly via google drive. The python code to analyze this data is available from this GitHub repository. The repository also includes code and instructions to reproduce fig. 4 from above images.

## Acknowledgements

Some of the illustrations shown in fig. 1, fig. 3, and fig. S3 were created with BioRender.com.

## Author contributions

J.R.B, E.W, O. J, S-M.G, J.V, and A.D.L designed, performed, and analyzed the experiments in supervision of B.G, J.E.P, C.M.T, and S.B.M. S-M.G, J.F, B.B.C, and S.B.M developed analytical pipeline. E.W, K.M.M, and J.E.P developed the ELISA-array, and J.E.P and W.W performed protein synthesis. H.L, I.E.I, and M.L.F designed and built the plate reader in supervision of M. P. and S.B.M., and collected data with it. M.J.P, T.J.H, S.G.D, and B.G organized the LIINC study and guided analysis. J.R.B, E.W, O.J, S-M.G, B.B.C, J.E.P, and S.B.M. wrote the paper with inputs from all authors.

## Supplementary Figures

**Fig. S1.**
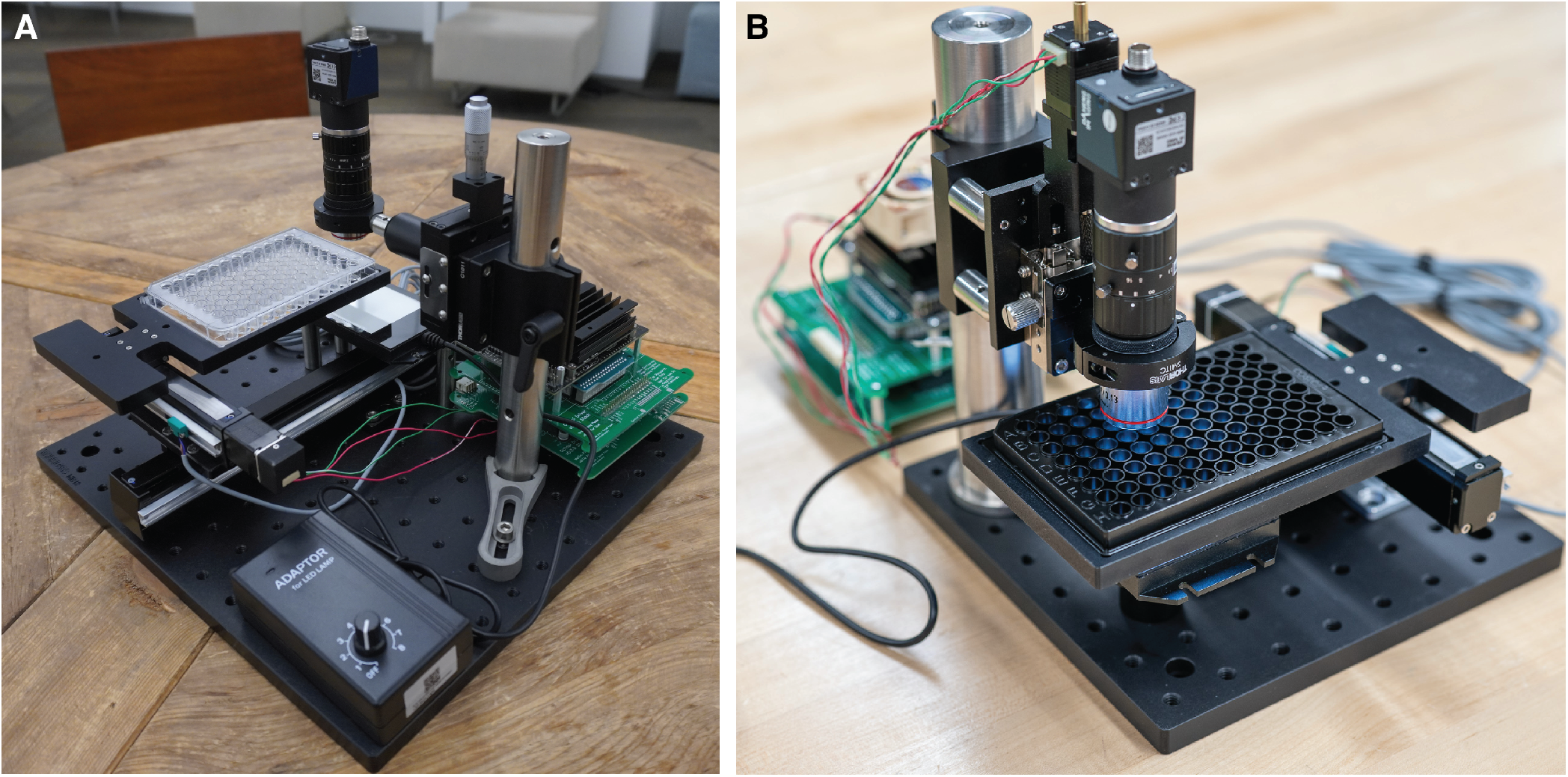
Photos of Squid plate reader (Nautilus). **(A)** Photo of the original unit used for data collection. **(B)** Photo of the current version that has motorized focus adjustment.

### Tables

**Table S1.**
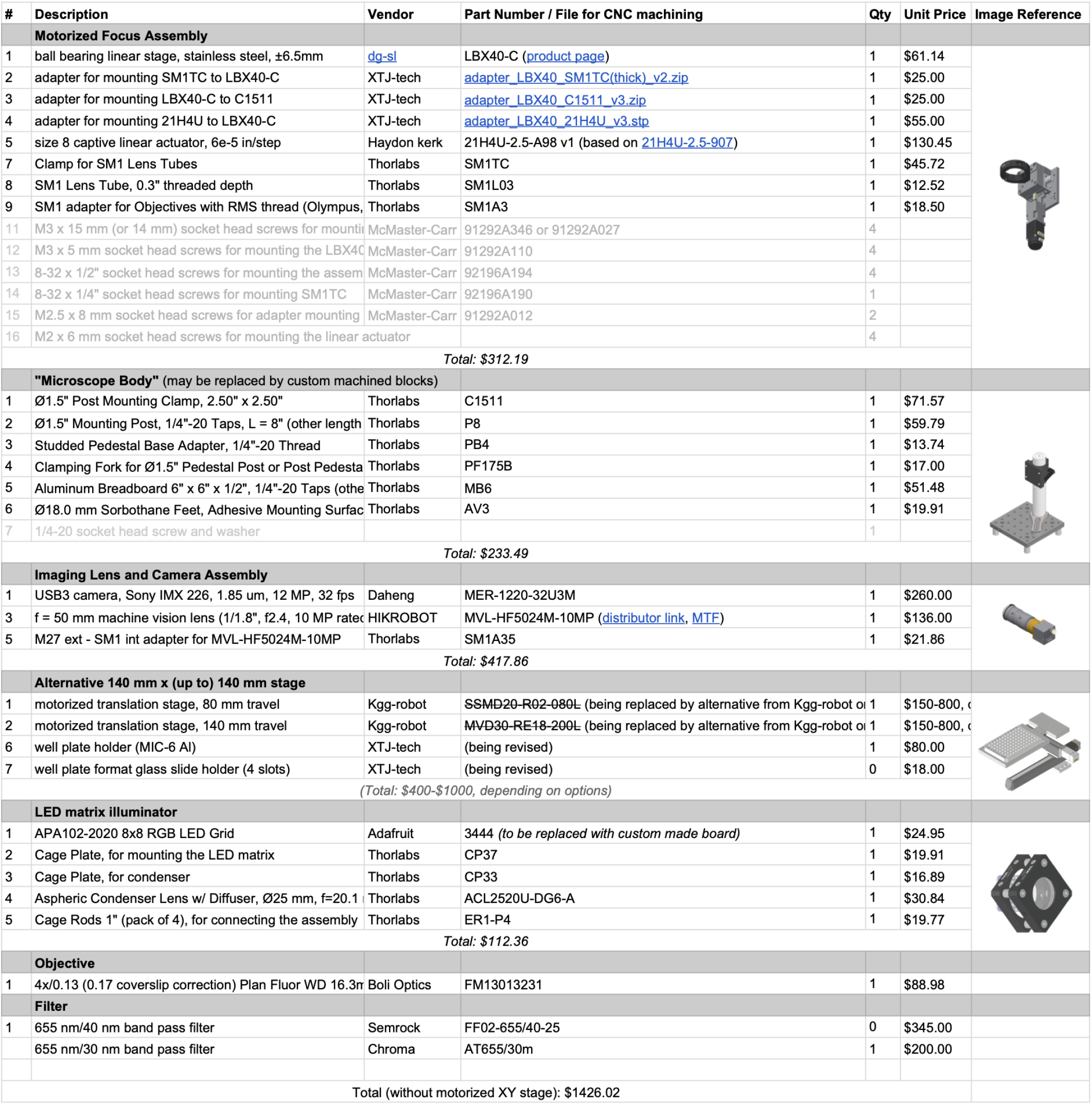
Nautilus Bill of Materials.

| # | Description | Vendor | Part Number / File for CNC machining | Qty | Unit Price | Image Reference |  |
| --- | --- | --- | --- | --- | --- | --- | --- |
| Motorized Focus Assembly |  |  |  |  |  |  |  |
| 1 | ball bearing linear stage, stainless steel, ±6.5mm | dg-sl | LBX40-C (product page) | 1 | \$61.14 | | |
| 2 | adapter for mounting SM1TC to LBX40-C | XTJ-tech | adapter_LBX40_SM1TC(thick)_v2.zip | 1 | \$25.00 | | |
| 3 | adapter for mounting LBX40-C to C1511 | XTJ-tech | adapter_LBX40_C1511_v3.zip | 1 | \$25.00 | | |
| 4 | adapter for mounting 21H4U to LBX40-C | XTJ-tech | adapter_LBX40_21H4U_v3.stp | 1 | \$55.00 | | |
| 5 | size 8 captive linear actuator, 6e-5 in/step | Haydon kerk | 21H4U-2.5-A98 v1 (based on 21H4U-2.5-907) | 1 | \$130.45 | | |
| 7 | Clamp for SM1 Lens Tubes | Thorlabs | SM1TC | 1 | \$45.72 | | |
| 8 | SM1 Lens Tube, 0.3" threaded depth | Thorlabs | SM1L03 | 1 | \$12.52 | | |
| 9 | SM1 adapter for Objectives with RMS thread (Olympus, Thorlabs | Thorlabs | SM1A3 | 1 | \$18.50 | | |
| 11 | M3 x 15 mm (or 14 mm) socket head screws for mounting the linear actuator | McMaster-Carr | 91292A346 or 91292A027 | 4 |  |  |  |
| 12 | M3 x 5 mm socket head screws for mounting the LBX40 | McMaster-Carr | 91292A110 | 4 |  |  |  |
| 13 | 8-32 x 1/2" socket head screws for mounting the assembly | McMaster-Carr | 92196A194 | 4 |  |  |  |
| 14 | 8-32 x 1/4" socket head screws for mounting SM1TC | McMaster-Carr | 92196A190 | 1 |  |  |  |
| 15 | M2.5 x 8 mm socket head screws for adapter mounting | McMaster-Carr | 91292A012 | 2 |  |  |  |
| 16 | M2 x 6 mm socket head screws for mounting the linear actuator |  |  | 4 |  |  |  |
| Total: \$312.19 | | | | | | | |
| "Microscope Body" (may be replaced by custom machined blocks) |  |  |  |  |  |  |  |
| 1 | Ø1.5" Post Mounting Clamp, 2.50" x 2.50" | Thorlabs | C1511 | 1 | \$71.57 | | |
| 2 | Ø1.5" Mounting Post, 1/4"-20 Taps, L = 8" (other lengths available) | Thorlabs | P8 | 1 | \$59.79 | | |
| 3 | Studded Pedestal Base Adapter, 1/4"-20 Thread | Thorlabs | PB4 | 1 | \$13.74 | | |
| 4 | Clamping Fork for Ø1.5" Pedestal Post or Post Pedestal | Thorlabs | PF175B | 1 | \$17.00 | | |
| 5 | Aluminum Breadboard 6" x 6" x 1/2", 1/4"-20 Taps (other sizes available) | Thorlabs | MB6 | 1 | \$51.48 | | |
| 6 | Ø18.0 mm Sorbothane Feet, Adhesive Mounting Surface | Thorlabs | AV3 | 1 | \$19.91 | | |
| 7 | 1/4-20 socket head screw and washer |  |  | 1 |  |  |  |
| Total: \$233.49 | | | | | | | |
| Imaging Lens and Camera Assembly |  |  |  |  |  |  |  |
| 1 | USB3 camera, Sony IMX 226, 1.85 µm, 12 MP, 32 fps | Daheng | MER-1220-32U3M | 1 | \$260.00 | | |
| 3 | f = 50 mm machine vision lens (1/1.8", f2.4, 10 MP rated) | HIKROBOT | MVL-HF5024M-10MP (distributor link, MTF) | 1 | \$136.00 | | |
| 5 | M27 ext - SM1 int adapter for MVL-HF5024M-10MP | Thorlabs | SM1A35 | 1 | \$21.86 | | |
| Total: \$417.86 | | | | | | | |
| Alternative 140 mm x (up to) 140 mm stage |  |  |  |  |  |  |  |
| 1 | motorized translation stage, 80 mm travel | Kgg-robot | SSMD20-R02-080L (being replaced by alternative from Kgg-robot or 140 mm travel) | 1 | \$150-800, contact for quote | | |
| 2 | motorized translation stage, 140 mm travel | Kgg-robot | MVD30-RE18-200L (being replaced by alternative from Kgg-robot or 140 mm travel) | 1 | \$150-800, contact for quote | | |
| 6 | well plate holder (MIC-6 Al) | XTJ-tech | (being revised) | 1 | \$80.00 | | |
| 7 | well plate format glass slide holder (4 slots) | XTJ-tech | (being revised) | 0 | \$18.00 | | |
| (Total: \$400-\$1000, depending on options) | | | | | | | |
| LED matrix illuminator |  |  |  |  |  |  |  |
| 1 | APA102-2020 8x8 RGB LED Grid | Adafruit | 3444 (to be replaced with custom made board) | 1 | \$24.95 | | |
| 2 | Cage Plate, for mounting the LED matrix | Thorlabs | CP37 | 1 | \$19.91 | | |
| 3 | Cage Plate, for condenser | Thorlabs | CP33 | 1 | \$16.89 | | |
| 4 | Aspheric Condenser Lens w/ Diffuser, Ø25 mm, f=20.1 | Thorlabs | ACL2520U-DG6-A | 1 | \$30.84 | | |
| 5 | Cage Rods 1" (pack of 4), for connecting the assembly | Thorlabs | ER1-P4 | 1 | \$19.77 | | |
| Total: \$112.36 | | | | | | | |
| Objective |  |  |  |  |  |  |  |
| 1 | 4x/0.13 (0.17 coverslip correction) Plan Fluor WD 16.3mm | Boli Optics | FM13013231 | 1 | \$88.98 | | |
| Filter |  |  |  |  |  |  |  |
| 1 | 655 nm/40 nm band pass filter | Semrock | FF02-655/40-25 | 0 | \$345.00 | | |
| | 655 nm/30 nm band pass filter | Chroma | AT655/30m | 1 | \$200.00 | | |
| Total (without motorized XY stage): \$1426.02 | | | | | | | |

**Fig. S2.**
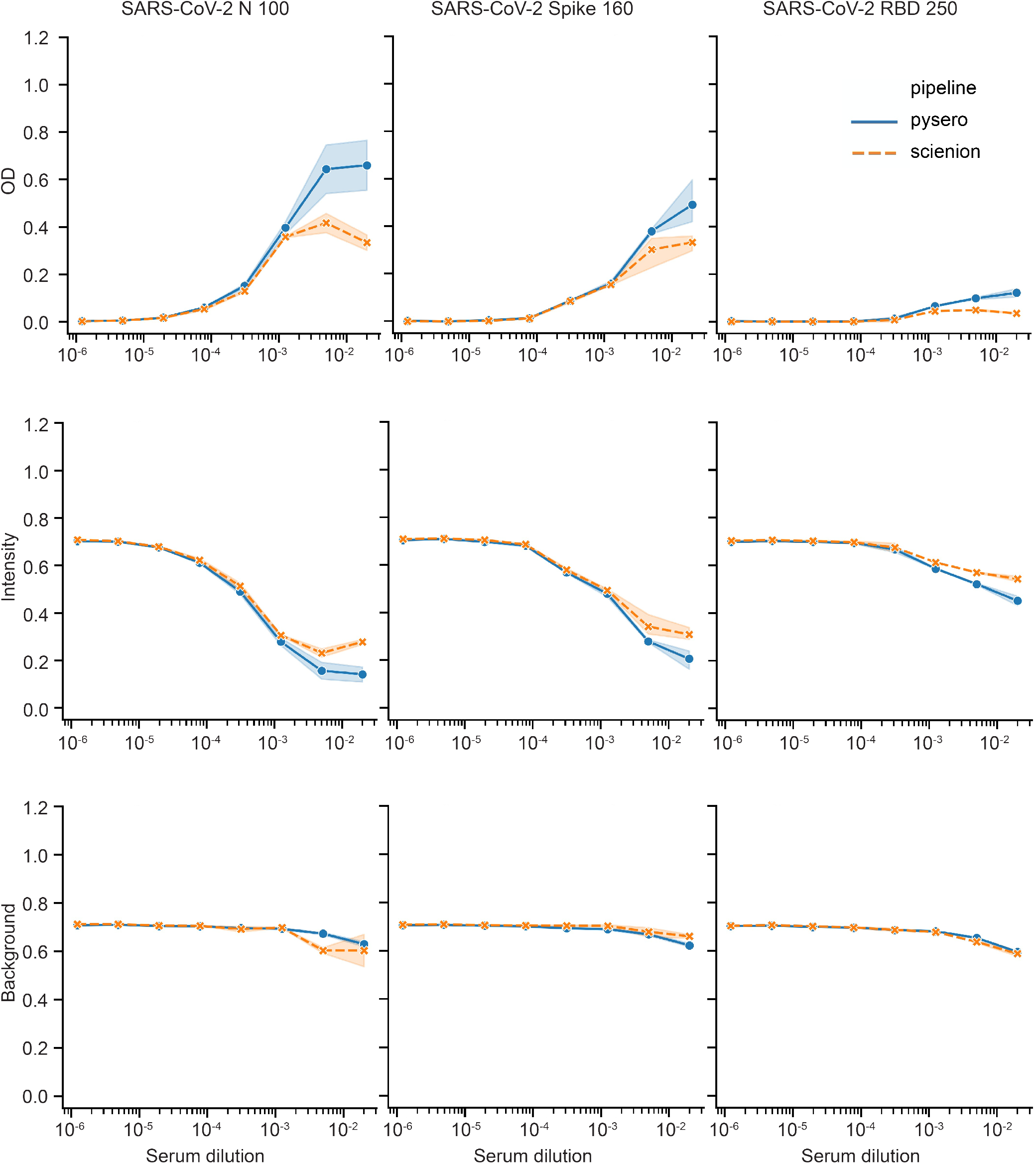
Comparison of Scienion analysis platform and pysero. Images from a SciREADER CL2 were analyzed using either pysero (blue line) or Scienion analysis platform (orange dashed line). Analyzed OD (top), intensity (middle), and background intensity (bottom) for antibody responses of a single SARS-CoV-2 positive serum to three antigens (left to right: SARS-CoV-2 N, Spike, RBD) are shown as a function of serum dilution. Shades around lines represent 95% confidence intervals around the mean of triplicate spots.

**Fig. S3.**
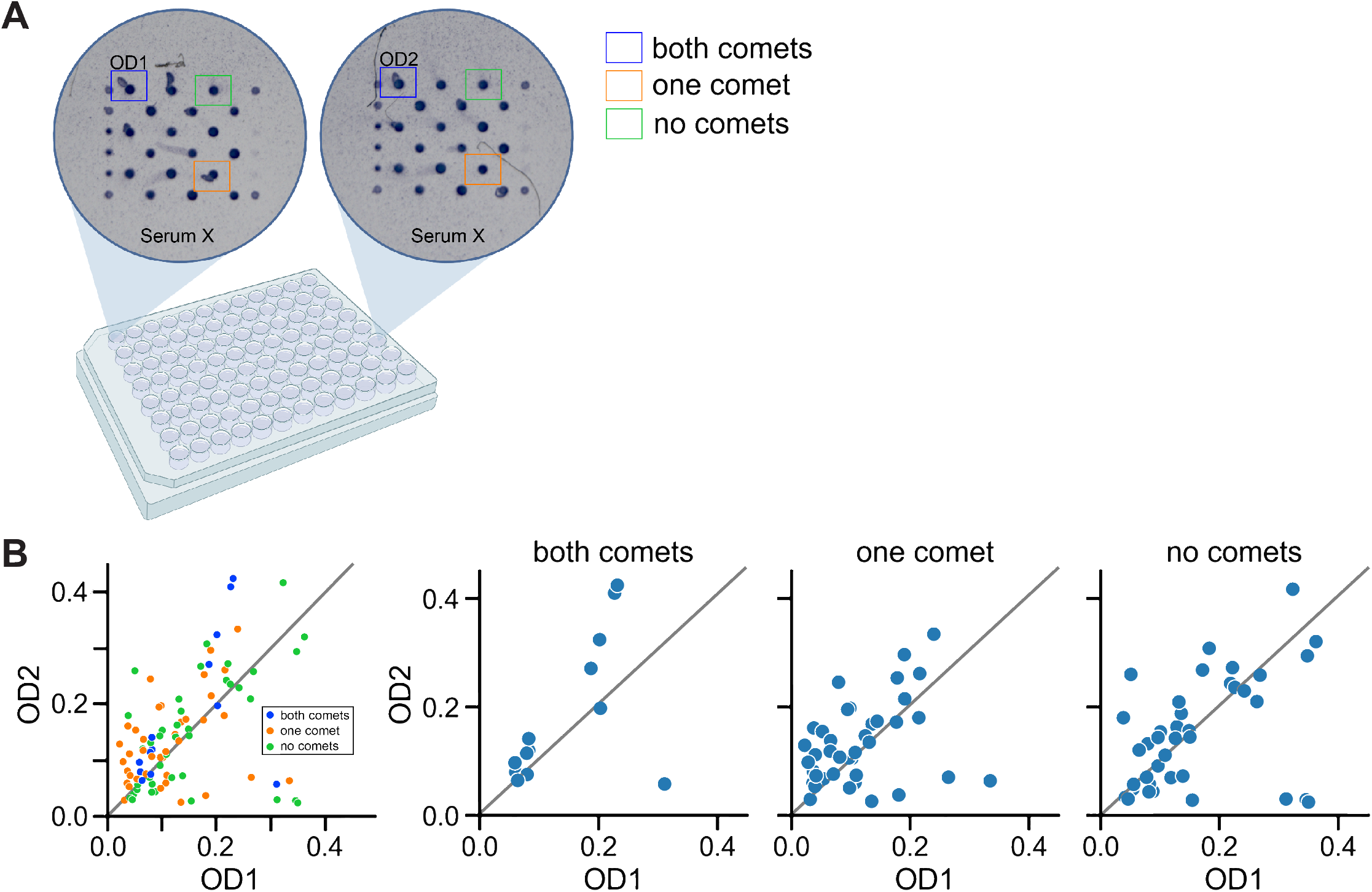
Evaluation of how comets affect measured ODs using duplicate ELISA-array wells. ODs from spots at the same locations in the array grid were compared across duplicate wells. **(A)** Schematic of example spot-spot comparison. Top-left non-fiducial spot in well A1 containing serum X was compared to the top-left non-fiducial spot in well F12 also containing serum X. **(B)**The data for one plate of duplicate sera are split according to the number of comets in the spot pairs: spot pairs in which one spot had a comet (orange); spot pairs in which both spots had comets (blue); and spot pairs in which neither spot had a comet (green). A y = x line is denoted on the plot (grey line). We find that the presence of comets does not add observable bias or variance to OD measurements.

**Fig. S4.**
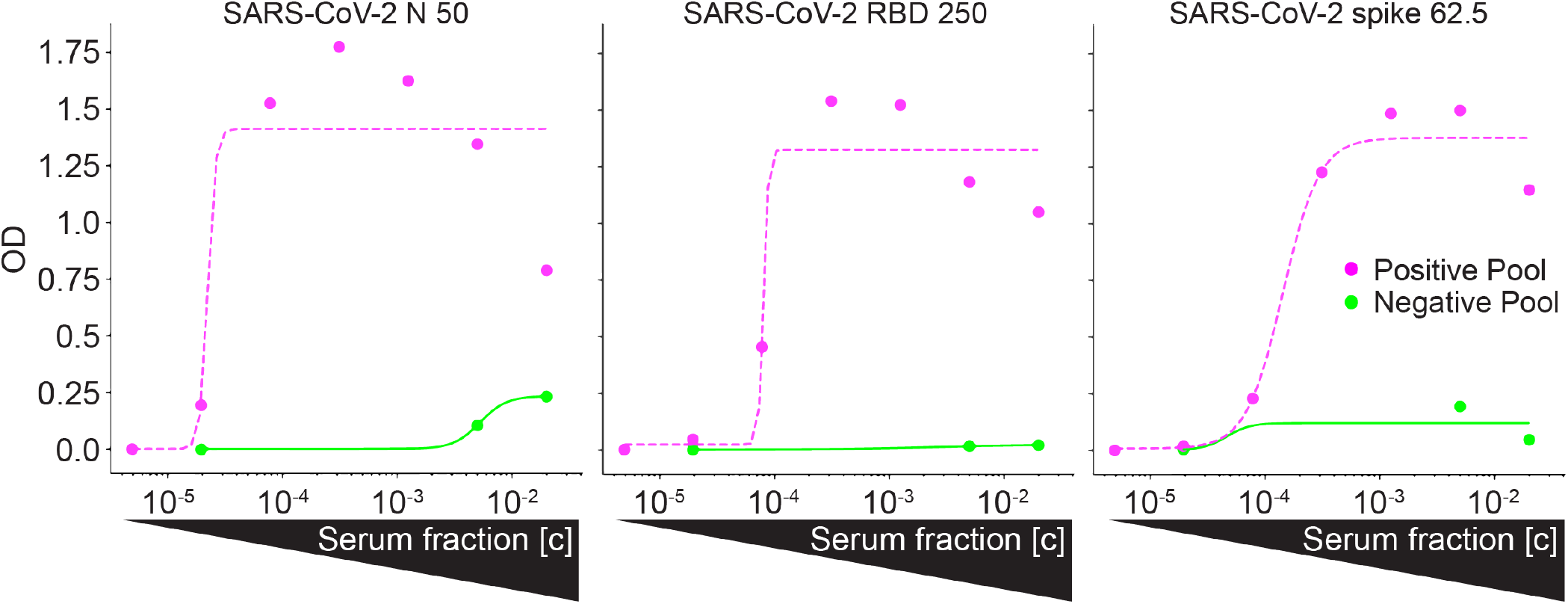
Reactivity of Positive Pool and Negative Pool. After OD extraction from the registered grid coordinates, pysero generates plots according to user-specified parameters. Representative plots of antibody responses against the 3 SARS-CoV-2 antigens in the array: N (left), RBD (center), and spike (right). Positive Pool indicates pooled sera from SARS-CoV-2 RT-PCR-positive individuals (magenta points) and Negative Pool indicates pooled sera from individuals prior to the pandemic (green points). The respective curves are parameter logistic regressions fit to OD values of a serial dilutions of the sera, with diminished OD values at highest concentrations likely due to the ‘hook effect’ (8). Pysero can output curve fits (shown above), categorical plots, and receiver operating characteristic analysis.

**Table S2.** Summary statistics for antigens in ELISA-array. Area under the curve (AUC), sensitivity (true positive rate; TPR) at 95% and 99% specificity (N=507), mean OD and mean binding of Negative Pool expressed as relative concentration of mAb CR3022 standard curve in Negative Pool ([Ab] of Neg Pool; units are µg mL*−1*) for each antigen. Asterisks in the relative antibody concentration of Negative Pool column indicate that the mean OD of the Negative Pool for these antigens was lower than the lowest mAb CR3022 concentration in the standard curve.

| Antigen | AUC | TPR 95% Specificity | TPR 99% Specificity | Mean +- sd OD of Neg Pool | Mean +- sd [Ab] of Neg Pool |
| --- | --- | --- | --- | --- | --- |
| N 50 | 0.969 | 0.907 (0.816-0.939) | 0.638 (0.398-0.826) | 0.1047 ± 0.02238 | 0.0011 ± 0.00055 |
| N 100 | 0.968 | 0.907 (0.822-0.943) | 0.541 (0.238-0.834) | 0.1087 ± 0.02432 | 0.0079 ± 0.00036 |
| RBD 250 | 0.983 | 0.977 (0.957-0.992) | 0.977 (0.936-0.992) | 0.0160 ± 0.01834 | 0.0004 ± 0.00025 |
| RBD 500 | 0.982 | 0.977 (0.957-0.992) | 0.977 (0.957-0.992) | 0.0230 ± 0.00735 | * |
| Spike 62.5 | 0.994 | 0.988 (0.973-1) | 0.984 (0.905-0.994) | 0.1933 ± 0.04399 | 0.0134 ± 0.00266 |
| Spike 125 | 0.989 | 0.984 (0.969-0.996) | 0.981 (0.897-0.992) | 0.0927 ± 0.01977 | * |

**Fig. S5.**
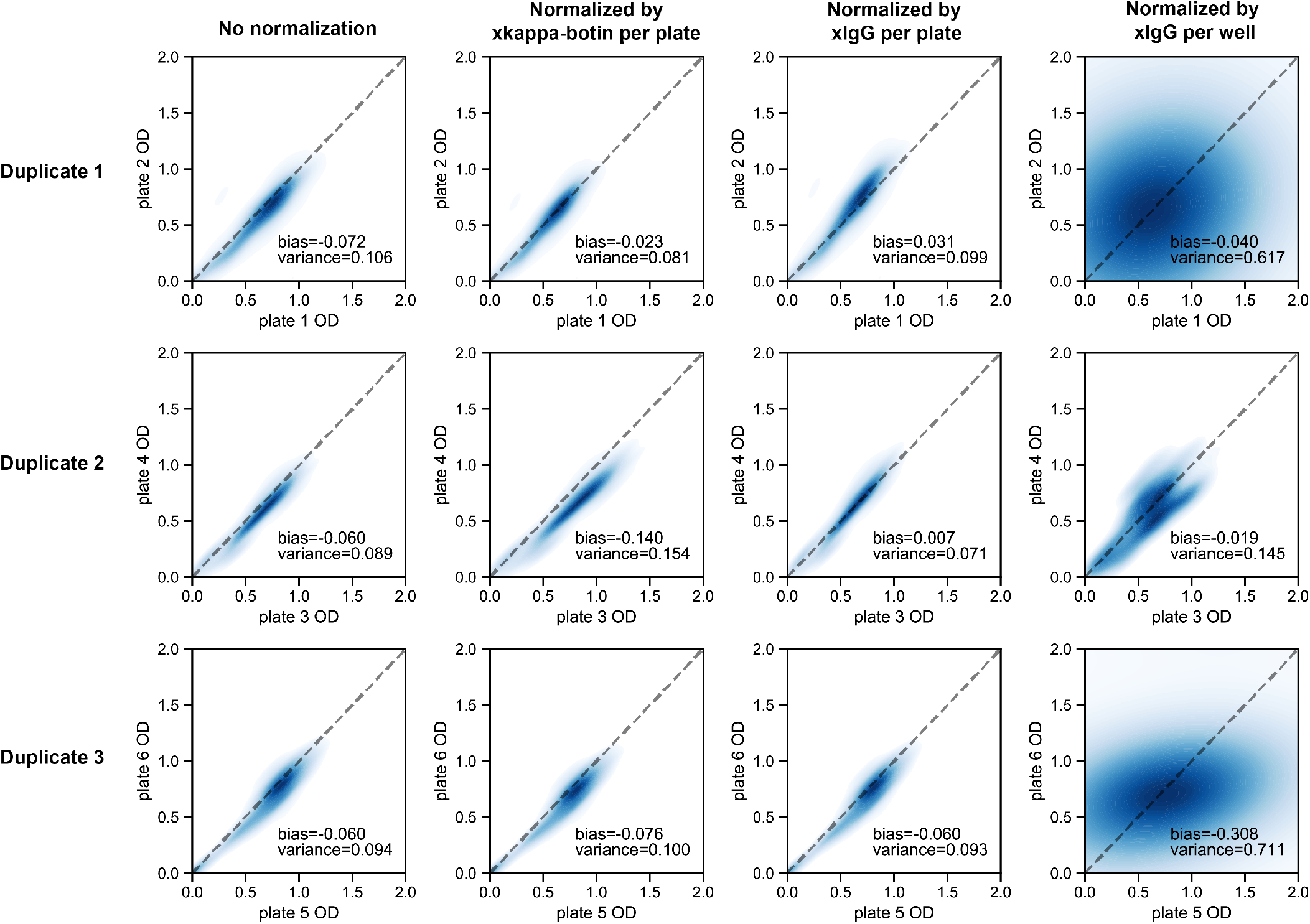
Comparison of normalization methods for correcting biases and variances in ODs across plates. 2D distribution of duplicate OD values of antibody responses of RT-PCR positive sera to SARS-CoV-2 antigens. Spots with the same array location on duplicate plates are plotted against each other and then smoothed by the kernel-density function to show the density of the data points (indicated by the brightness of the blue color, high brightness indicates low density). The spot OD values were normalized by dividing the mean of the reference spot ODs (anti-IgG Fc or fiducial spot) over each plate or well as indicated at the top. Duplicates with identical spot OD values will follow the function y = x (dashed line). Performance of different normalization schemes are quantified by bias and variance of the normalized OD values across plates, which are defined by mean of y – x and |y – x|. 3 duplicates (6 plates) are shown. Normalizing ODs by the mean of anti-IgG Fc ODs over each plate consistently reduces bias and variance in all 3 duplicates.

**Fig. S6.**
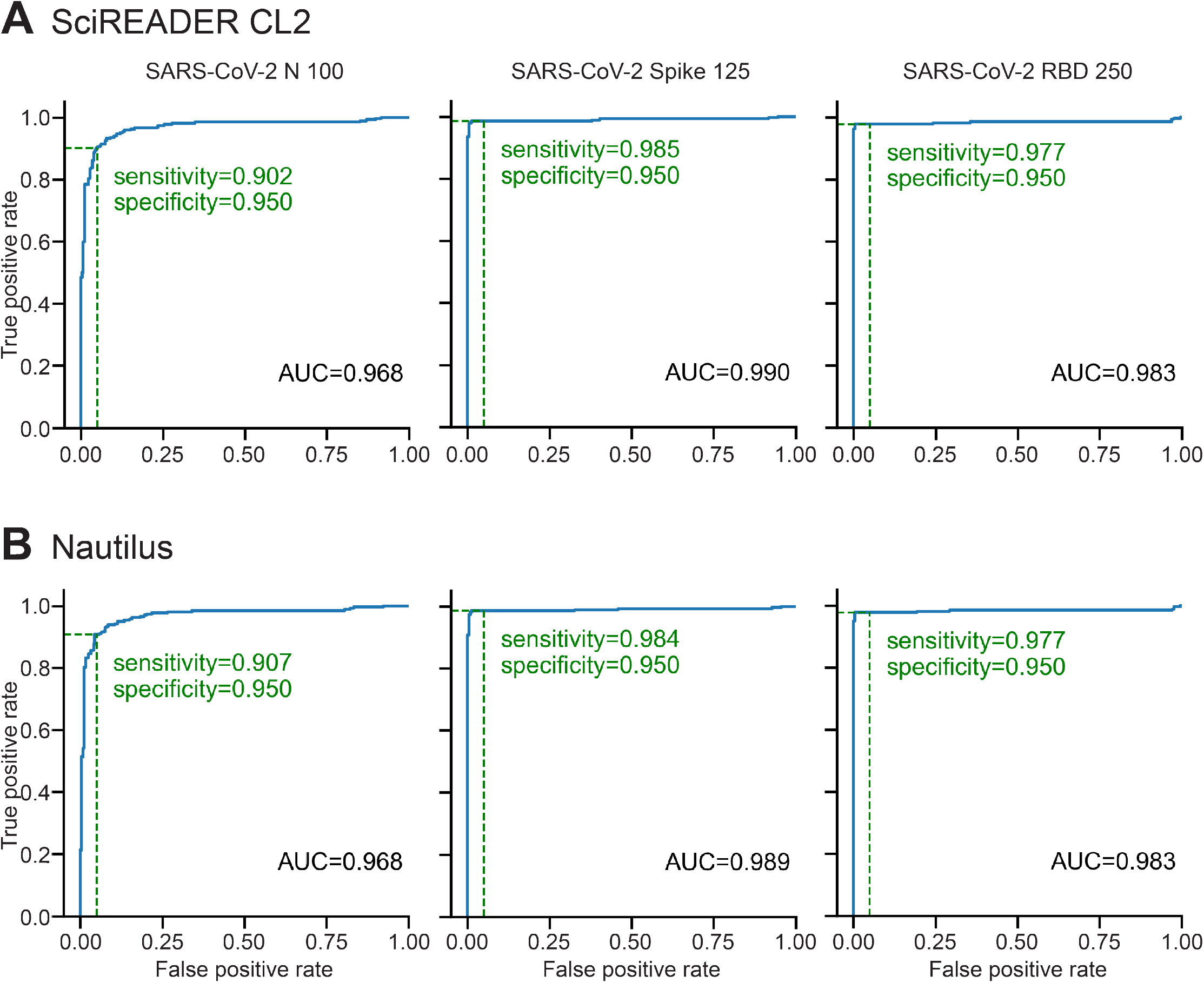
Classification accuracy for images acquired with SciREADER CL2 and Nautilus, and analyzed with pysero. **(A-B)** ROC curves for single antigens (left to right: SARS-CoV-2 N, Spike, RBD) for array images acquired using SciREADER CL2 (top) or Nautilus (bottom). Images from each platform were analyzed using pysero. Blue shades represent 95% confidence intervals of ROC curves (blue line). For each antigen, the sensitivity (True positive rate) at 95% specificity (1 - false positive rate) is denoted (green dashed line) on the curve. 95% confidence interval of area under the curve (AUC) is reported below each curve (N=507).

